# Understanding Diagnostic Error Patterns and Contributing Factors: A Descriptive Analysis of Medical Error Reports at a Tertiary Hospital in Kenya 2019-2021

**DOI:** 10.1101/2024.05.21.24307687

**Authors:** Lydia Okutoyi, Pamela Godia, Mary Adam, Fred Sitati, Walter Jaoko

## Abstract

**Background:** Diagnostic errors in healthcare pose substantial risks, leading to increased costs, patient anxiety, and delayed diagnoses. Despite its prevalence, diagnostic errors have historically received less attention compared to other medical errors, necessitating urgent action to address these critical issues. This is more so in the low- and middle-income countries. (LMICs). This study aimed to analyze patterns and associated factors of diagnostic error reported to the Patient Safety Unit of Kenyatta National Hospital (KNH), a tertiary teaching hospital in Nairobi, Kenya.

**Methods:** This was a descriptive retrospective study of medical error reports(MER) forms submitted to KNH from 2019-2021.Type of medical errors, contributing factors, site, timing of error, and outcome were recorded. Descriptive statistics, chi-square tests, and logistic regression were employed to assess error types, contributing factors, and associated likelihoods.

**Results:** Among 640 MER forms analysed, diagnostic errors were reported in 40 percent of cases, predominantly associated with delayed diagnosis, wrong diagnosis, and failure to test. Contributing factors to MER included communication issues (36.1%), staff-related factors (48.9%), and equipment issues (15.6%). Diagnostic errors were more likely during non-working hours (OR 1.969, p < 0.047) and in Accident and Emergency department (OR 2.36, p < 0.022) within KNH.

**Conclusion:** Diagnostic errors represent a significant proportion of medical errors at KNH, particularly in Accident and Emergency settings. Strategies to involve more physicians in error reporting and enhance communication practices are recommended.

## INTRODUCTION

Diagnostic errors pose significant risks and challenges in healthcare delivery, often leading to increased costs, heightened patient anxiety, and, in critical cases, delayed diagnoses [1, 2]. Research indicates that these errors are pervasive, with one in every 20 patients experiencing such an error in population-based studies [3]. Diagnostic errors are noted to have a likelihood of causing moderate to severe harm compared to other types of errors[4]. Despite their prevalence, diagnostic errors have historically received less attention compared to other medical errors, necessitating urgent action to address these critical issues [5–7]. This discrepancy has underscored the urgent need for focused research and interventions to address diagnostic errors comprehensively.

Diagnostic error, broadly defined as missed opportunities in diagnosis or follow-up actions based on available evidence, reflects both provider and systemic shortcomings [1]. Errors can manifest as delayed, incorrect, or missed diagnoses. Research by Graber et al. (2005) and Henriksen et al. (2015) has highlighted the complexities of diagnostic errors, emphasizing the multifaceted nature of these incidents[6, 8]. The errors can arise from a variety of causes, including cognitive biases, system failures, and breakdowns in communication [9, 10].

Addressing diagnostic errors requires robust strategies for identification and documentation to enable accurate measurement. Previous reliance on autopsy and malpractice reports has proven inadequate, highlighting the need for a systemic approach that extends beyond individual practitioners to encompass the entire diagnostic process [6, 8]. Indeed being able to measure diagnostic errors is essential. Despite increasing awareness of diagnostic errors, they remain challenging to identify and report within healthcare systems [4]. Healthcare providers involved in diagnostic errors often exhibit reluctance to report incidents due to concerns about professional reputation and legal repercussions [4, 5]. This reluctance highlights the critical need for improved reporting mechanisms that prioritize learning and system improvement over punitive measures.

Diagnostic errors pose reporting challenges across various medical specialties, including instances occurring in operating theatres or identified through radiology examinations [6, 11]. Medical error reporting systems, while essential for patient safety, often present incomplete pictures of incidents due to the timing of reporting and the inherent limitations of individual clinical judgment [3, 11]. In the USA, diagnostic safety has been accorded the need attention by the academics and the clinician in the annual Diagnostic Errors in Medicine (DEM) conferences over the past decade, yet this topic remains less addressed in low- and middle-income countries (LMIC) [5].

Patient-provider interactions were the highest contributing factors for diagnostic errors, while patient related factors were the lowest in a Japan study where patients who had an unscheduled visit back to the outpatient department within 14 days of prior visit[12]. In a large study among 21 hospitals in the Netherlands human failure (96.3%) was identified as the main cause of diagnostic adverse events, while organizational and patient related factors also contributed (25% and 30.0% respectively)[13]. Other studies note patient-provider factors and communication among the care team have been noted to be the two leading contributors to diagnostic errors[14, 15].

Despite the existence of a Medical Error Reporting (MER) system at Kenyatta National Hospital (KNH) for seven years, detailed studies on diagnostic errors within this framework are lacking. This study aimed to analyze the patterns and factors associated with diagnostic errors reports.

## METHODS

### Study Design and Setting

This was a descriptive retrospective study. Data were collected from medical error reports submitted to the Kenyatta National Hospital (KNH) Medical Error Reporting System spanning from January 2019 to December 2021. Data extraction and data entry were conducted between January 2022 and March 2022. Kenyatta National Hospital is a tertiary care teaching hospital in Nairobi, Kenya. During this period, KNH utilized a paper-based Medical Error Reporting (MER) form. The form is a 14-field document that adheres to World Health Organization (WHO) standards for reporting medical errors while maintaining patient and healthcare provider anonymity[16]. It includes key fields such as the date of error occurrence, patient information (excluding identifiers like names), and error classification into diagnostic, treatment, medication, or preventive categories. It also captures a brief description of the error, its impact on patients or processes, contributing factors (e.g., human or system-related), and details of actions taken for mitigation and prevention. The reporting officer fills out the initial 13 fields, and the unit leader, often a nurse, completes the final sections to ensure comprehensive documentation. Physical copies of filled MER forms were submitted to the Patient Safety Unit every month.

### Study Population and Sampling

All MER forms submitted during the study period were included in the analysis. All KNH clinical departments are encouraged to identify medical errors, document and submit filled MER forms to the patient safety unit monthly. Summary reports generated within the hospital and respective departments which didn’t have individual forms were not included in the study.

### Data Collection Tool Development and Training

A data abstraction tool was developed in REDCap, aligned with the World Health Organization (WHO) minimum information tool for MER[16]. Five research assistants (RAs) who were statistics and medical students from local universities, were selected and trained to use the online data collection tool. They familiarized themselves with MER forms and were supervised by the Principal Investigator (PI). The research assistants were supported to pilot at least 5 forms each to ensure data accuracy during the first week of MER form abstraction.

### Data Collection Process

RAs entered data in a designated room within the healthcare quality department, adhering to confidentiality and privacy guidelines. Clarifications on abbreviations, job roles, diseases, and clinical processes were provided using a glossary of commonly used terminologies at KNH. Data entry involved extracting information directly from the filled fields in the MER forms. Parts of the MER forms included short narratives describing what happened and the outcomes, while other sections featured checkboxes. In cases where data was missing, we utilized the brief narratives provided in the MER forms to construct certain fields. Throughout this process, the principal investigator and staff from the patient safety unit provided guidance to the research assistants.

### Variables of Interest

The variables collected were the type of medical errors reported (diagnostic errors, treatment errors, medication errors, preventive errors, or other types), location (Accident and Emergency, Ward and other), Time (normal working hours-8:00am to 5:00pm weekdays, and non-normal working hours), Contributing factors to error happening (Communication, Patient related, Staff related, Equipment related and Lack of policies).

### Data Analysis

Descriptive analysis was conducted to summarize error types and subtypes. Continuous variables were presented as mean ± standard deviation (SD), while categorical variables were reported as numbers and percentages. Chi-square test was used for inter-group comparisons of quantitative variables, with p-values <0.05 considered statistically significant.

A multivariate logistic regression model was employed to analyse associations between diagnostic error types, time of error occurrence, and whether errors occurred within KNH. The model was adjusted, and odds ratios with a 95% confidence interval were calculated. McFadden’s R2 was used to assess model fit.

Data were analysed using Jamovi 2.3.21 and RStudio 4.2.2 which are open-source for statistical computing.

### Ethical consideration

The study received ethics approval from the Kenyatta National Hospital-University of Nairobi Ethics Review Committee (Approval No. P847/10/2021) and obtained a research permit from the National Commission for Science Technology and Innovation (Permit Ref. 517313). The department involved authorized data collection using stored forms, with the Head of the unit signing an institutional research form. No identifiable information was collected from the forms, and no human samples or experiments were conducted as part of this research.

## RESULTS

Six hundred and forty (640) medical error report forms were submitted to the Patient Safety Unit at the Kenyatta National Hospital. Age, diagnosis, and gender had 16.7%, 12.3%, and 6.5% missing data, while the rest of the variables of interest had less than 5% missing data.

### Characteristics for patient’s, medical errors, and location of reporting

The distribution of medical errors in relation to gender, age, diagnoses, reporting personnel and their outcomes is shown in Table 1. There was a similar proportion between diagnostic and non-diagnostic errors, with males accounting for slightly more errors overall (54.2% of diagnostic errors vs. 52.9% of non-diagnostic errors). However, this difference was not statistically significant (p = 0.406).

**Table 1.** Distribution of medical errors in relation to gender, age, diagnoses, reporting personnel and their outcomes.

| Patient Characteristics |  |  | Diagnostics | Nondiagnosics | p-value |
| --- | --- | --- | --- | --- | --- |
|  |  | Totals |  |  |  |
| Gender |  |  |  |  |  |
|  | Female | 274(45.8%) | 105 | 169 | 0.406 |
|  | Male | 324(54.2%) | 135 | 189 |  |
| Age |  |  |  |  |  |
|  | 0-19 | 65(12.3%) | 21 | 44 | 0.054 |
|  | 20-39 | 241(45.5%) | 98 | 143 |  |
|  | 40-59 | 155(29.3%) | 72 | 83 |  |
|  | 60-79 | 53(10.0%) | 22 | 31 |  |
|  | 80-99 | 16(3.0%) | 2 | 14 |  |
| Diagnosis |  |  |  |  |  |
|  | Surgical | 324(57.6%) | 127 | 197 | 0.655 |
|  | Medical | 190(33.8%) | 82 | 108 |  |
|  | Oncology | 49(8.7%) | 19 | 30 |  |
| Person reporting |  |  |  |  |  |
|  | Doctor | 3(0.5%) | 0 | 3 | 0.185 |
|  | Nurses | 614(99.2%) | 248 | 366 |  |
|  | Pharmacy | 2(0.3%) | 0 | 2 |  |
| Outcomes |  |  |  |  |  |
|  | Good | 539(84.2%) | 210 | 329 | 0.215 |
|  | Poor | 101(15.8%) | 46 | 55 |  |

Although there was a higher proportion of errors occurring in the age group of 40-59 years for both diagnostic and non-diagnostic errors, the distribution between the errors with age was not statistically significant (Chi-square test; p=0.054).

Surgical cases accounted for most errors in diagnostic and non-diagnostic categories (57.6% vs. 58.0%), followed by medical and oncology cases. No significant differences were observed in the distribution of error types across diagnoses (p = 0.655).

Nurses were responsible for most error reports (99.2% overall), with minimal contributions from doctors and pharmacy staff. The difference in reporting personnel was not statistically significant (p = 0.185). Most reported errors had good outcomes (84.2% overall), with a slightly higher proportion of poor outcomes associated with diagnostic errors (15.8%) compared to non-diagnostic errors (14.1%). However, this difference was not statistically significant (p = 0.215).

### Location of reporting and Time period of error happening

The location of reporting and time of error happening in the hospital is shown in Table 2. Accident and Emergency (AE) had the highest proportion (45.5%) of diagnostic errors among the three areas, followed by Wards (38.2%) and Other (31.9%). Other locations included Theatre, Critical care units, and Pharmacy. The observed proportion of diagnostic errors was higher during non-normal working hours (46.1%) compared to normal working hours (35.1%).(Table 2)

**Table 2.**
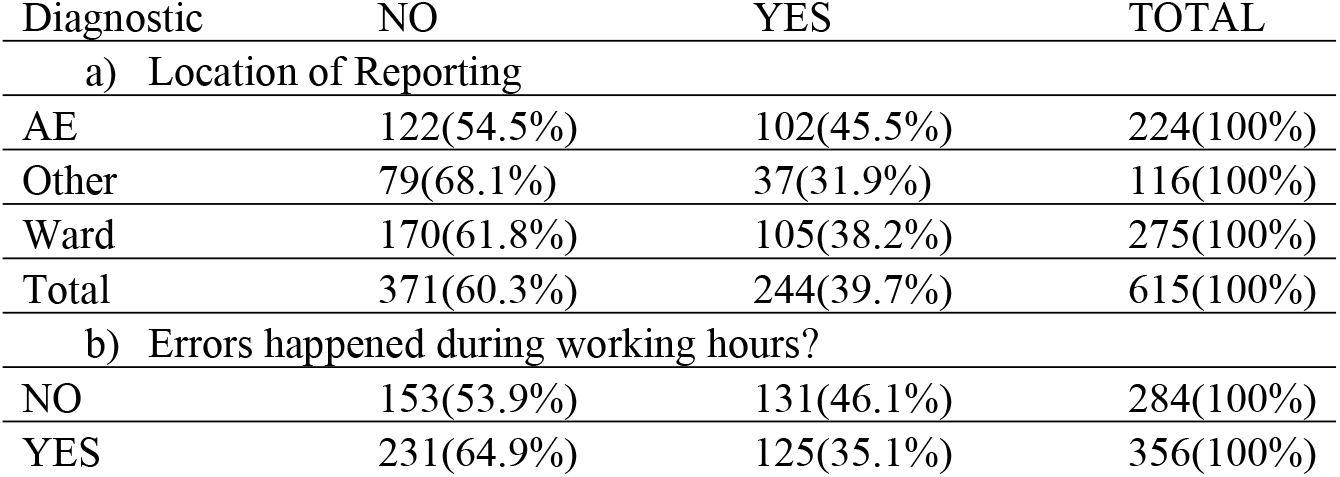
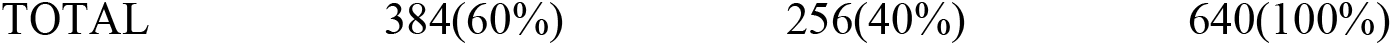
Distribution by of error reporting in relation to location and time error happened.

### Types of Medical Errors reported

The types of reported errors are shown in Table 3. Diagnostic errors and treatment errors emerged as the most frequently reported types of medical errors. Diagnostic errors were identified in 40% (256 forms) of the submissions. Treatment-related errors were the most frequently reported, comprising 54.2% (347 forms) of the submissions, followed by medication errors at 17.8% (114 forms). Other identified error categories included prevention (7.0%, 45 forms), documentation (16.3%, 104 forms), and miscellaneous errors classified as ‘Others’ (20.3%, 130 forms).

**Table 3;.**
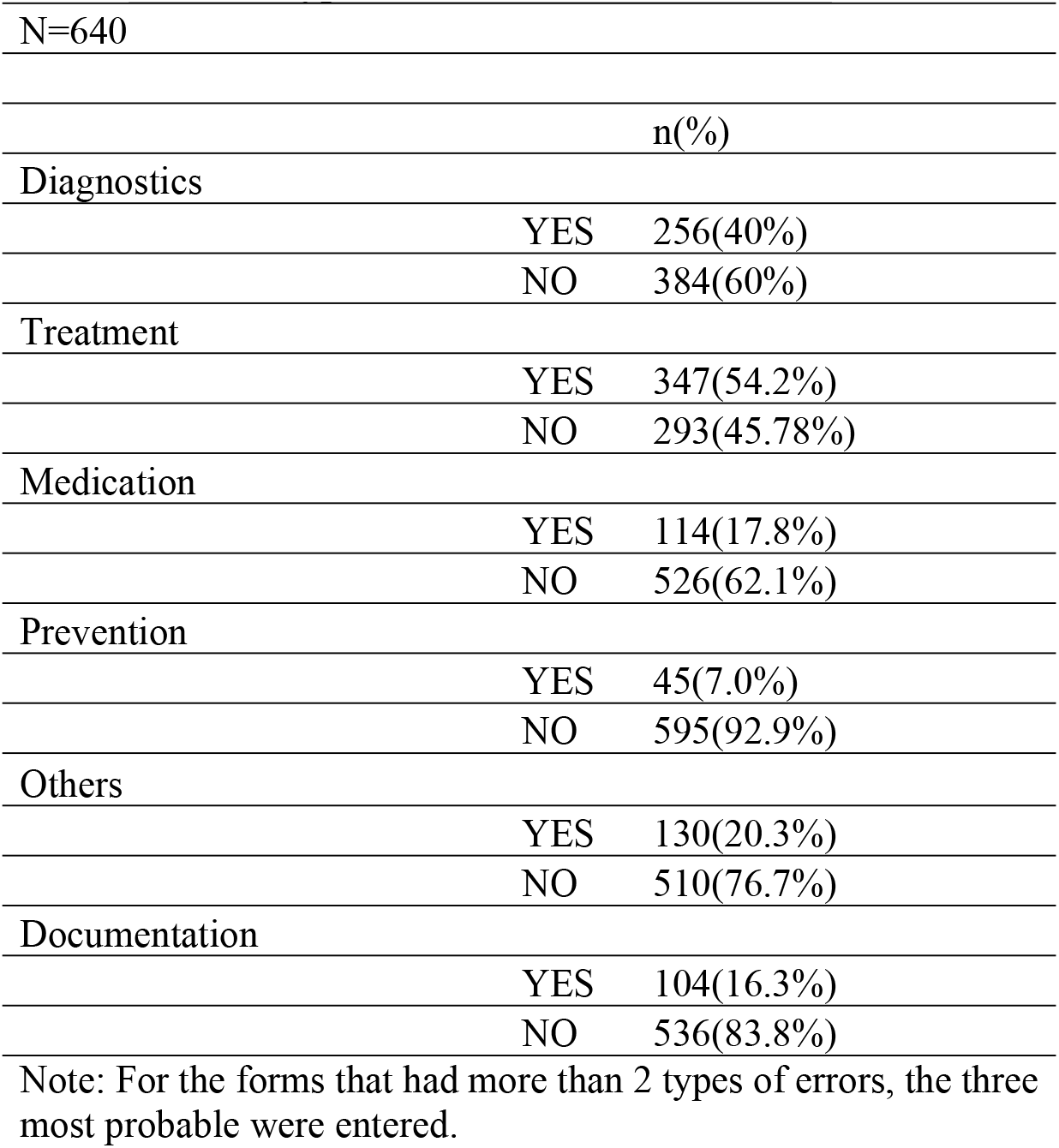
Type of Medical errors submitted.

Of the 256 reported diagnostic errors, the top three subtypes of errors reported were: delay in diagnosis (45.9% of diagnostic errors), wrong diagnosis (27.2%), and failure to carry out required test (8.7%). See Figure 1.

**Figure 1;.**
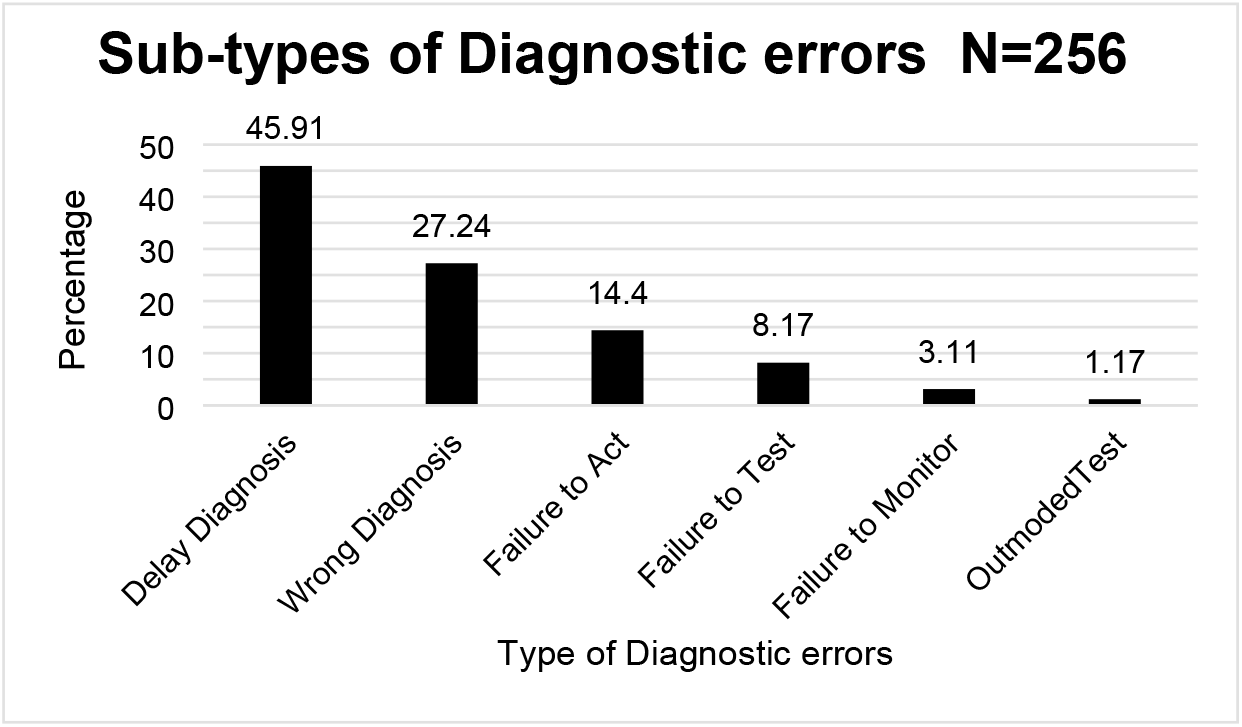
Subtypes of diagnostic errors

For the forms that had more than 2 types of errors, the three probable were entered.

### Contributing Factors to Medical Errors: Diagnostic vs. Non-Diagnostic Errors

Various factors contributed to the occurrence of medical errors as shown on table 4. These included communication issues, accounting for 23.1% of cases, patient-related factors at 11.6%, staff-related issues at 48.9%, equipment failures or deficiencies at 15.6%, and deficiencies in policies which contributed to 21.4% of the total 640 medical errors.

**Table 4;.** Contributing factors to diagnostic errors.

|  |  | Totals | Diagnosis Errors | Nondiagnostic Errors | p values |
| --- | --- | --- | --- | --- | --- |
| Communication |  |  |  |  |  |
|  | YES | 231(36.1%) | 101 | 130 | 0.149 |
|  | NO | 409(63.9%) | 155 | 254 |  |
| Patient factors |  |  |  |  |  |
|  | YES | 74(11.6%) | 34 | 40 | 0.26 |
|  | NO | 566(88.4%) | 222 | 344 |  |
| Staff factors |  |  |  |  |  |
|  | YES | 313(48.9%) | 126 | 187 | 0.89 |
|  | NO | 327(51.1%) | 130 | 197 |  |
| Equipment |  |  |  |  |  |
|  | YES | 100(15.6%) | 35 | 65 | 0.27 |
|  | NO | 540(84.4%) | 221 | 319 |  |
| Lack of Policies/ guidelines |  |  |  |  |  |
|  | YES | 137(21.4%) | 64 | 73 | 0.07 |
|  | NO | 503(78.6%) | 192 | 311 |  |

**Table 5:** Associations of factors with the likelihood of reporting diagnostic errors. (Reference level; Diagnostic-NO, Normal working hours reference-YES, Location KNH reference NO, Location of reporting -Ward).

**Model Coefficients - Diagnostic**
|  |  |  |  |  |  | 95% Confidence Interval |  |
| --- | --- | --- | --- | --- | --- | --- | --- |
| Predictor | Estimate | SE | Z | P | Odds ratio | Lower | Upper |
| Intercept | -1.49 | 0.332 | -4.5 | <.001 | 0.225 | 0.118 | 0.431 |
| Normal Working Hours: |  |  |  |  |  |  |  |
| NO – YES | 0.341 | 0.172 | 1.98 | 0.047 | 1.406 | 1.004 | 1.969 |
| Location of Reporting: |  |  |  |  |  |  |  |
| AE – Ward | 0.463 | 0.202 | 2.29 | 0.022 | 1.589 | 1.069 | 2.362 |
| Other – Ward | -0.284 | 0.236 | -1.2 | 0.228 | 0.753 | 0.475 | 1.195 |
| Location of Error KNH?: |  |  |  |  |  |  |  |
| YES – NO | 0.894 | 0.301 | 2.96 | 0.003 | 2.444 | 1.354 | 4.412 |

Note. Estimates represent the log odds of "Diagnostic = YES" vs. "Diagnostic = NO"
R<sup>2</sup>McF 0.0235

Communication issues were linked to 101 diagnostic errors and 130 non-diagnostic errors, revealing no statistically significant difference (p = 0.26) in their influence on error occurrence. Out of the 74 medical errors attributed to patient-related factors, 34 were diagnostic errors and 40 were non-diagnostic errors, with no statistically significant difference observed in their occurrence between the groups.

Staff-related factors were associated with 126 out of 313 diagnostic errors and 187 out of 313 non-diagnostic errors, with no statistically significant difference observed (p = 0.89) between the rates of diagnostic and non-diagnostic errors related to staff factors.

For equipment issues, 35 diagnostic errors and 65 non-diagnostic errors were attributed to lack or faulty equipment, with no statistically significant difference in their contribution to error occurrence (p = 0.27).

Regarding the absence of policies or guidelines, 64 out of 137 diagnostic errors and 73 out of 417 non-diagnostic errors were influenced by this factor, showing a trend towards significance (p = 0.07) in the difference between diagnostic and non-diagnostic error rates related to policy deficiencies.

### Associations of factors with the likelihood of reporting diagnostic errors

The odds of diagnostic error happening during the nonworking hours was 1.969 times that of it happening during normal working hours OR 1.969 (95% CI:1.004, 1.406) and was statistically significant at (p <0.047).

The odds of diagnostic error being reported in the Accident and Emergency unit was 2.36 times that of it being reported in the wards OR 2.36 (95% CI: 1.069, 1.589) and was statistically significant at (p <0.022).

The odds of diagnostic error happening in KNH was 1.195 times that of it happening outside KNH OR 1.195 (95% CI:1.354, 2.444) and was statistically significant at (p <0.003). Note that KNH is a tertiary institution that receives referral from lower level facilities from within the environs of Nairobi and country Kenya.

## DISCUSSION

Diagnostic errors are the second most common medical errors submitted to the hospital MER system, following treatment errors. The Accident and Emergency department had a higher likelihood of reporting diagnostic errors compared to the wards, and more reports of diagnostic errors occurred during non-working hours than during normal working hours.

### The incidence of diagnostic errors

Diagnostic errors account for a significant portion (40%) of reported errors at KNH. A study in Uganda by Katongole, found Diagnostic errors being the commonest errors among 618 charts abstracted and accounted for 40.5% of all the ME, the same study also found Healthcare Workers (HCW) stated the perceived frequency of medication errors (58%) and diagnostic errors (53%)[17]. This finding are similar to the incidence in this study, likely because of similar setting, though the Uganda study was not from a voluntary reporting system. In a primary care setting in the UK Aveery et al found ; 60.8% of the adverse events reported were diagnostic errors. This study by Aveery etal had a small sample size of 74, likely explaining the high proportions[18]. However, a review of a nationwide incidence reporting in the same country between 2003-2005 indicated a lower incidence at 0.5% [4].

A review of studies in the USA found the incidence of diagnostic errors in the hospital setting to be between 6.4%-17% of the adverse events[1]. The difference can be explained by the larger sample size.

#### Patient characteristics

The commonest age group in this study was 20-39 years, which comprised 45% of the forms, while the male to female ratio was 1:1.2. Aoki et al, did a patient reported study on diagnostic errors in Japan, where the commonest age group was 60-79 years (61%), while this study had a younger age group as the most frequent [19]. The study in Japan was an outpatient study and also the population in Japan having much older population than that in Kenya where this study was done. Gende distribution in this study in fairly comparable to that in the Japan study; male: female ration 1:1.2 [19].

Patients with oncology diagnosis made 8.75%, surgical 57.6%, and medical 33.8%. Oncology patients in the study by Aoki et al. were similar at 8.3%, while the medical and Surgical was 47.2% and 43.7% respectively[19]. Patients with medical diagnosis were more in the study by Aoki because of this being a primary care setting, while the surgical diagnosis was higher in our study because the hospital provides emergency care services.

The vast majority (99.2%) of forms submitted to the system were from nurses. Research shows that nurses consistently report a higher percentage of errors (67%-93%) compared to doctors (2%-23%) in incident reporting systems. Even when comparing the proportion of nurses reporting to their total number versus doctors reporting to the total number of doctors, nurses still have a higher percentage reporting. At KNH, the nursing department has made medical error reporting a performance target, which has increased awareness of MER compared to the doctors.

The likelihood of diagnostic errors happening during non-working hours were 1.96 more than during the normal working hours. A study in the Saudia Arabia established that medication errors were more likely to happen at night compared to the day duty[21]. This is similar to this study, human factor challenges are more apparent at night than normal hours.

### Diagnostic errors at Accident and Emergency

In this study A&E had the highest proportion (45.5%) of diagnostic errors among the three areas, followed by Ward (38.2%) and Other (31.9%). Evaluating the diagnostic errors in the National UK reporting system, diagnostic incidents were thus less likely to occur in a ward (P < 0.002) and more likely to occur in an emergency department (P < 0.002) than other incidents[4]. The higher percentage of diagnostic errors in A&E suggests a higher likelihood more errors happening during transitions and primary care.

### Contributing factors of errors happening

Staff factors(48%), Communication(36%) and lack of policies(21%) were the three leading contributing factors to medical errors occurring in this study. While reviewing surgical medical-legal cases in a Canadian study, provider and communication factors contributed to 80% and 50% of the cases respectively[14]. A multicentre study among 71 Dutch hospitals also identified human error and communication among the teams as the common contributing factors to diagnostic errors[15]. In deed this is similar to our study where Staff related issues and communication topped the list.

### Study limitation

The study’s scope encompasses a retrospective analysis of medical error reports at KNH over three years, focusing on diagnostic errors. Limitations include potential underreporting and incomplete data within the MER system. Some of the fields had multiple types of medical errors and contributing factors, which were handled by data cleaning, this has been described in the methods section.

## CONCLUSION

Diagnostic errors are the second most common medical errors submitted to the hospital MER system, following treatment errors.. Accident and emergency is an important area of focus for identification of diagnostic errors from other levels of care and also those errors made during emergency care. This is the first study to describe the patterns in the KNH medical error reporting system. Strategies of involving more doctors in documentation of medical error using the MER form should be implemented. Research using chart abstraction and patient reported diagnostic errors is likely to offer more information on the prevalence of diagnostic error, and shade light on the diagnostic process than this descriptive study.

## Data Availability

Data is available is needed. This does not have patient identifiers and is available for sharing.

## Notes

### Competing Interest Statement

The authors have declared no competing interest.

### Funding Statement

This study was funded by the Kenyatta National Hospital for the purpose of quality improvement in diagnostic safety.

### Author Declarations

The study received ethics approval from the Kenyatta National Hospital-University of Nairobi Ethics Review Committee (Approval No. P847/10/2021) and obtained a research permit from the National Commission for Science Technology and Innovation (Permit Ref. 517313).

